# Reading; through the eyes of a university student: A double-masked randomised placebo-controlled cross-over protocol investigating coloured spectacle lens efficacy in adults with visual stress

**DOI:** 10.1101/2024.08.18.24312188

**Authors:** Darragh L. Harkin, Julie-Anne Little, Sara J. McCullough

## Abstract

**Background:** Visual stress is a reading disorder characterised by perceptual distortions, asthenopia and headache whilst reading, alongside increased sensitivity to repeated striped patterns (‘pattern glare’), in the absence of underlying ocular pathology. Coloured filters including tinted spectacle lenses and coloured overlays/acetates have been reported to ameliorate visual stress symptoms. However, evidence on coloured spectacle lenses efficacy at managing symptoms of visual stress, particularly in adults, is lacking, with two recent systematic reviews advocating the need for large-scale randomised control trials.

**Methods:** This is a double-masked randomised placebo-controlled trial. University students identified with symptoms of visual stress, through use of a reading symptom questionnaire and mid-spatial frequency pattern glare test, will be recruited. Sample size for power of 90% at 5% significance, accounting for 10% dropout will be 65. Participants will be randomly assigned experimental and control coloured spectacle lenses to wear for six weeks followed by a two week washout period, prior to wear of the alternate lenses for a further six weeks with a two week washout period. Participants will compare both sets of spectacle lenses in a ‘head-to-head’ comparison after the secondary washout period, prior to choosing the preferred lenses for voluntary future wear. Long-term adherence to the preferred lenses will be assessed three months post-comparison. Researchers and participants will be masked to spectacle lenses worn throughout the duration of the trial. Reading performance will be assessed with both sets of lenses at various time points within the trial. A range of reading tests, reading symptoms and pattern glare evaluation will be used to monitor change in reading performance and visual stress symptoms during the trial.

**Discussion:** The study will evaluate the hypothesis that coloured spectacle lenses increase reading speed and reduce severity and frequency of reading symptoms in adults with visual stress.

**Trial Registration:** The trial is registered at clinicaltrials.gov: NCT04318106

## Introduction

Visual Stress, occasionally referred to as ‘Meares-Irlen Syndrome’ and ‘Scotopic Sensitivity Syndrome’ is a visuoperceptual reading disorder with a reported prevalence varying between 13-96% [1] in both adults and children. Variable prevalence can be explained by the array of symptoms and signs employed to diagnose the condition, sample population tested [2][3] and unclear evidence on the underlying aetiology. In an absence of standardised diagnostic criteria [3][4], the condition is diagnosed by eyecare practitioners on an ad-hoc basis. Attempts have been made to standardise diagnostic criteria [3][5][6][7] however these have not been validated. Colorimetry – the practice of determining preferred chromaticity of coloured spectacle lenses to assist with reading symptoms, is performed in some primary eyecare settings. Coloured filters, in the form of tinted spectacle lenses or sheets of coloured acetate have been reported to improve reading performance and reduce symptoms in individuals with visual stress [6][8][9][10] in small-scale randomised controlled trials and case-controlled studies.

Symptoms of visual stress can be broadly described as the presence of perceptual distortions, such as words and letters ‘moving’ or ‘merging’, and appearance of ‘rivers’ or coloured patterns within lines of crowded text; typically black text printed on white backgrounds [3][7]. There are several aetiologies proposed to cause visual stress but, in brief, these either fall under (i) A deficit in magnocellular processing and inability to process high temporal frequency ‘flicker’ [11][12] or (ii) Hyperexcitation of the visual cortex from repetitive black-on-white striped patterns often found in written text [13][14]. With regard to theory (i), coloured spectacle lenses are proposed to reduce the extent of cortical hyperexcitation by attenuating the amount of potentially aversive visual information that is received by a hyperexcitable primary visual cortex [15][16]. For theory (ii), coloured filters are thought to supress potentially aberrant intraretinal image patterns on saccadic eye movements [11][17] and improve ocular movements whilst reading [18] in symptomatic individuals with underlying magnocellular pathway deficits.

Whilst the theories underpinning the aetiological mechanisms of visual stress are well explained in the literature, investigations into the efficacy of coloured lenses at ameliorating symptoms of visual stress are scant. Two recent systematic reviews, both conclude that a large-scale double-masked randomised control trial needs to take place to investigate coloured lens efficacy in subjects with visual stress [1][19]. Additionally, professional optometric bodies’ management guidance in relation to the practice of colorimetry is guarded, citing a lack of robust evidence to either endorse or reject the practice [20][21][22]. This protocol will detail the methods employed in a double-masked cross over randomised placebo-controlled trial. We aim to test the hypothesis that providing coloured lenses to those with symptoms of visual stress will be effective at improving reading performance and reduce frequency of symptoms experienced whilst reading.

## Methods & Materials

### Study Design

This study is designed as a double-masked cross-over randomised placebo-controlled trial of coloured lens efficacy in adults with symptoms of visual stress. The trial design is supported by the Standard Protocol Items: Recommendations for Intervention Trials (SPIRIT) framework (see supporting information S1). Participants will be evenly and randomly assigned one of two sets of coloured spectacle lenses for initial spectacle wear for a period of six weeks, followed by wear of an alternate pair of spectacle lenses for a further six weeks. Coloured spectacle lenses will differ in their chromaticity, with the ‘experimental’ coloured lenses, tinted to optimal chromaticity, which are presumed to reduce perceptual distortion symptoms, whilst ‘control’ lenses will comprise a chromaticity similar in perceptual colour appearance to ‘experimental’ lenses, to assist with masking, but differing in chromaticity, such that visual stress symptoms are neither improved nor worsened with use of tint. The ‘Control’ lenses shall therefore act as a placebo. Researchers and participants will both be masked to the spectacle lenses worn. A two week washout between periods of spectacle lens wear will be employed, so that any potential legacy effects of coloured lenses are negated.

### Study Population

#### Sample Size Calculation

The Wilkins Rate of Reading Test reading speed has been used as the outcome measure for an improvement with use of coloured filters [9][10][23][25][26]. A within-subject standard deviation of 10.85 words per minute has been calculated from estimated values from repeat measures data on 77 school-age children [27]. The expected difference between the two treatments (i.e. ‘control’ lenses vs ‘experimental’ lenses) has been estimated as 6.5 words per minute, derived from data on a non-dyslexic population for adults with visual stress [6]. A sample size of 59 participants was determined appropriate for this investigation with a power of 90% and an alpha level of 0.05. The required sample size has been increased by 10% to account for attrition. Therefore, 65 participants shall be recruited for the trial.

#### Study Location & Participant Demographics

Participants will be recruited from the undergraduate student population at Ulster University, Coleraine, Northern Ireland, UK and all study visits will take place at the Centre for Optometry and Vision Science at Ulster University.

#### Inclusion Criteria

All participants shall be adult undergraduate students at Ulster University, aged 18-45-years. Participants shall be recruited in their first year of their current university degree. Participants will be enrolled in the trial on the basis of presence of symptoms suggestive of visual stress, as determined by a reading symptom questionnaire and as detailed in Table 1

#### Exclusion Criteria

Participants will be excluded if they satisfy any one of the following criteria:

- Habitual distance monocular acuity poorer than 0.1 logMAR.
- Habitual near monocular acuity poorer than N5 at 40cm.
- Presence of manifest strabismus and/or decompensating phoria observed on alternating cover test.
- Near point of convergence less than 10cm.
- Failure to meet Duane-Hoffstetter Criteria for minimum binocular push-up accommodative amplitude (>15-[0.25]*[age in years]) – see Monger et al.[28]
- Absence of observable stereopsis on the TNO stereotest.
- Formal diagnosis of photosensitive epilepsy

### Recruitment

#### Reading Symptoms Screening

A reading symptom questionnaire was developed to screen large cohorts of undergraduate students in lecture theatres (see Supporting Information 2). There are currently no validated criteria used to diagnose visual stress [1][3]. Therefore, reading symptoms employed in the questionnaire were devised from symptoms of visual stress reported in the literature such as those proposed by a Delphi study [3] and the ‘Visual Stress Index’ [7]. The questionnaire includes 17-items and 6-point Likert scale with descriptors of symptom frequency (0=never; 5=always). Participants will be provided with a sample of crowded text in a foreign language, devoid of context, to consider when answering the questionnaire [7].

Further questions exploring participants’ ocular and general health will be posed, as well as questions investigating presence of potential visual stress co-morbidities such as; headache [29][30]; formal diagnosis of migraine[31][32][33][34]; diagnosis of dyslexia [6][35][36] and/or diagnosis of specific learning difficulties and other general health complaints, known to also be managed with coloured filters [16][37][38][39].

To determine presence of cortical hyperexcitability, a mid-spatial frequency pattern glare test will also be administered. A 40cm string will be affixed to the test, to measure the proposed viewing distance of ‘pattern 2’ of the pattern glare test series (spatial frequency 2.3cpd) [40]. Respondents will be directed to circle ‘yes/no’ responses to each of the seven pattern glare symptoms as per the test instructions [40][41]. Each positive ‘yes’ response will be summed to determine a total pattern glare score ranging from 0-7.

Reading symptom questionnaires will be administered during term time. The researchers will attend lecture theatres and inform students that they are conducting an investigation of symptoms students experience whilst reading. Recruitment to this study began on 08/12/2022. It is anticipated that final recruitment will be completed by 20/12/24. Participants will be informed that completion of the questionnaire is voluntary and that all information disclosed will be treated confidentially. Participants will not be informed of the potential to participate in the randomised control trial until after completion of the questionnaire to avoid biasing results.

#### Determination of the Presence of Visual Stress

In the absence of standardised diagnostic criteria for visual stress, recruitment criteria were devised on the basis of previously reported criteria and methods of recruitment to previous investigations within the literature [6][8][9]. One drawback of previous investigations of coloured filter efficacy is the inability of some studies to accurately determine presence of visual stress in participants, with some incorrectly enrolling participants on the basis of dyslexia diagnosis [42][43][44] or due to variations in definition of visual stress symptoms (see review by Griffiths et. al. 2016)[1]. The current study will employ strict recruitment criteria to ensure that the most symptomatic individuals are included in the study. Participants who satisfy at least one set of the inclusion criteria as listed in Table 1 shall be invited to take part in the trial. By satisfying either criterion, participants will be recruited either on the basis of (i) presence of pattern glare and at least two significant visuoperceptual reading symptoms indicative of visual stress or (ii) overall summed symptom severity across an array of 14-visuoperceptual symptoms.

**Table 1.**
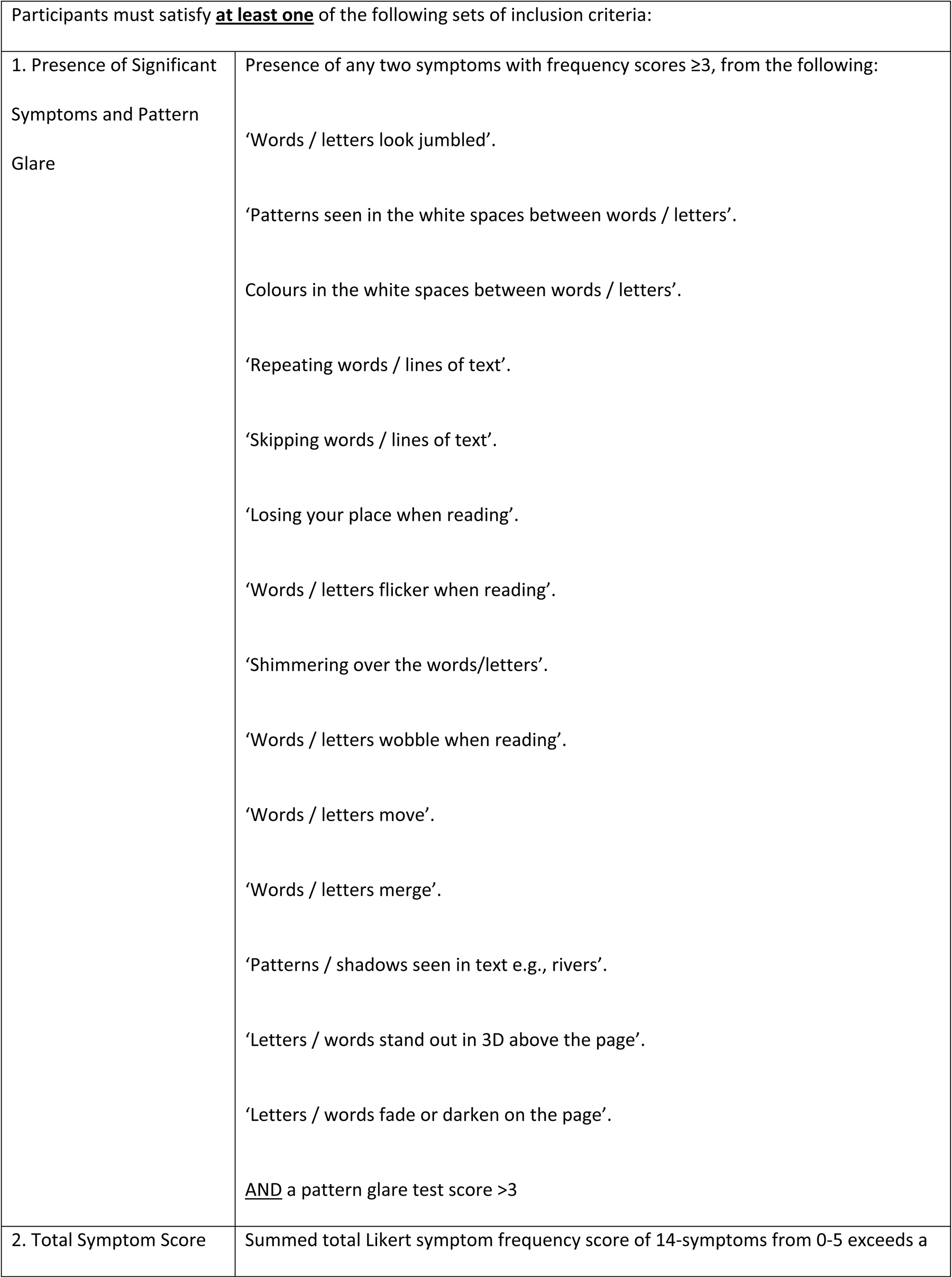
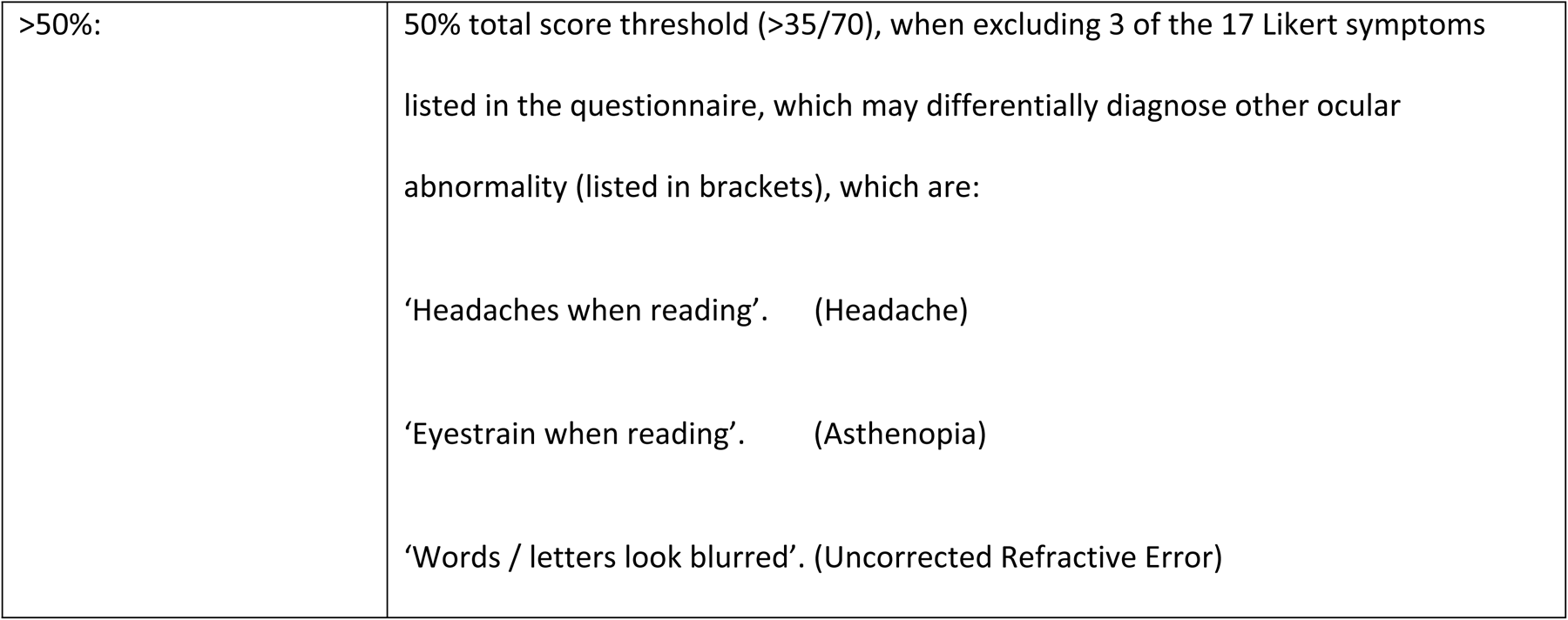
Recruitment Criteria used to Determine Presence of Visual Stress.

|  |  |
| --- | --- |
| 1. Presence of Significant Symptoms and Pattern Glare | Presence of any two symptoms with frequency scores $\geq 3$, from the following:<br>‘Words / letters look jumbled’.<br>‘Patterns seen in the white spaces between words / letters’.<br>Colours in the white spaces between words / letters’.<br>‘Repeating words / lines of text’.<br>‘Skipping words / lines of text’.<br>‘Losing your place when reading’.<br>‘Words / letters flicker when reading’.<br>‘Shimmering over the words/letters’.<br>‘Words / letters wobble when reading’.<br>‘Words / letters move’.<br>‘Words / letters merge’.<br>‘Patterns / shadows seen in text e.g., rivers’.<br>‘Letters / words stand out in 3D above the page’.<br>‘Letters / words fade or darken on the page’.<br><u>AND</u> a pattern glare test score $> 3$ |
| 2. Total Symptom Score | Summed total Likert symptom frequency score of 14-symptoms from 0-5 exceeds a |
| >50%: | 50% total score threshold (>35/70), when excluding 3 of the 17 Likert symptoms listed in the questionnaire, which may differentially diagnose other ocular abnormality (listed in brackets), which are:<br>‘Headaches when reading’. (Headache)<br>‘Eyestrain when reading’. (Asthenopia)<br>‘Words / letters look blurred’. (Uncorrected Refractive Error) |

### Procedure

#### Interventions Employed (Determination of Control Lens)

The ‘Curve’ Intuitive Colorimeter ™ [45] will be used to determine chromaticity of ‘experimental’ (optimal) and ‘control’ (sub-optimal) tints, through manipulation of the hue, saturation and luminance of a projected series of three primary colours to comprise a gamut of visible chromaticities spaced throughout the 1976 CIELUV colour space[46][47].

Participants will identify an ‘experimental’ tint through a series of repeated presentations of opposing hues. Initially, 12-predetermined hues, separated by a ‘hue angle’ of 30° will be presented. Participants will compare the appearance and comfort of a random series of letters forming a striped body of text devoid of context within the Intuitive Colorimeter™, illuminated under white light and presented hue. Participants will note if the presented hue makes the appearance and comfort of the crowded text ‘better’, ‘worse’ or ‘no-different’ than white-light. Beneficial hues will be shortlisted. Saturation of each shortlisted hue will be reduced until symptoms arise. Saturation will then be adjusted to the lowest level where symptoms resolve. A 2-alternative forced choice of preferred hue of ideal saturation will then be presented until an optimal chromaticity can be determined, as previously described [48]. Once optimal hue and saturation are determined, luminance will be attenuated between ‘half’ and ‘full’ settings and the participant offered a preference choice. An arc of beneficial therapeutic colours will be uncovered through this technique. This area of therapeutic chromaticities will represent an area on the 1976 CIELUV colour space wherein observers are expected to derive benefit from colour. The authors propose that variation in size and location of this therapeutic region will exist amongst observers. Boundaries of the colour space will be denoted as two ‘hue angles’ on the Intuitive Colorimeter™ colour wheel, outside which therapeutic benefit is lost. Optimal chromaticity will lie within this colour space. This arc of beneficial chromaticities will be herein referred to as the ‘Limit of Therapeutic Effect’.

The ‘control’ lens will act as the placebo and will be sub-optimal, when compared to ‘experimental’ tint. To determine control tint, researchers will prescribe a colour that will not be aversive, yet not beneficial to the patient’s symptoms of visual stress that lies just outside the ‘Limit of Therapeutic Effect’. The researchers will choose the chromaticity closest to the optimal to assist with masking of experimental and control lenses. ‘Control’ tint will be of the same saturation and luminance to the ‘experimental’ tint. Researchers are mindful of previous work, stating interpractitioner and inter-instrumental measurement error may account for ∼0.03 units of chromaticity difference (the Euclidean distance between colour coordinates mapped on the 1976 CIELUV Chromaticity Diagram) between lenses [49][50]. Whilst a consistent chromatic separation between lenses will not be applied, to account for assumed difference in size of ‘Limit of Therapeutic Effect’ by observers, size of chromatic separation between lenses will be considered in post-hoc analysis of reading performance.

The chromaticity difference, between colour coordinates of ‘experimental’ and ‘control’ lenses will be measured by the Mark 2 SpectroCAL Spectroradiometer (Cambridge Research Systems Ltd., Cambridge, UK). Chromaticity coordinates will be plotted by computer software (Light Values Measuring System, Version 6.2.0, Windows 11.0 OS, Jeti Technische Instrumente GmbH Tatzendpromenade 2, Jena, Germany). Chromaticity difference between ‘experimental’ and ‘control’ tints will be considered in post-hoc analysis of reading performance with the use of coloured lenses.

#### Randomisation & Blinding

Randomisation of interventions will be conducted by the Ulster University School of Biomedical Sciences’ Clinical Trials Manager. Researchers and participants will be blinded as to which lenses are assigned throughout the duration of the trial. Both spectacle frames housing the spectacle lenses will be identical to mask researchers and participants to the intervention employed.

#### Spectacle Wear Periods & Washout

Participants will wear each intervention for a period of six weeks. Participants will be informed that spectacle wear is voluntary and to wear as much as desired. Participants will be advised to avoid wearing coloured spectacle lenses when driving at night as per manufacturer’s specification [47], to ensure that spectral transmission complies with College of Optometrists’ Guidance for Professional Practice on provision of coloured filters when driving [51]. Within the first and final week of spectacle wear, participants will be asked to complete a spectacle wear diary online (Research Electronic Data Capture, version 11.0.3, 2021, or paper copy where necessary) at the end of each day of wear. The diary will record the number of hours the spectacles are worn daily. If the participant indicates that they have not worn the spectacles, they will be asked to provide reasons for this. Participants will also be asked to note the number of hours completing concentrated and/or near-work tasks and if the coloured spectacles have been worn to assist with these tasks. Participants will also be asked to grade how helpful (if at all), the spectacle lenses have been in assisting their reading performance on a six point Likert scale (0 = ‘Not at all helpful’ to 5 = ‘Extremely helpful’). Fig. 2 comprises a flowchart of the questions that will be posed to participants in their online spectacle wear diary.

**Figure 1.**
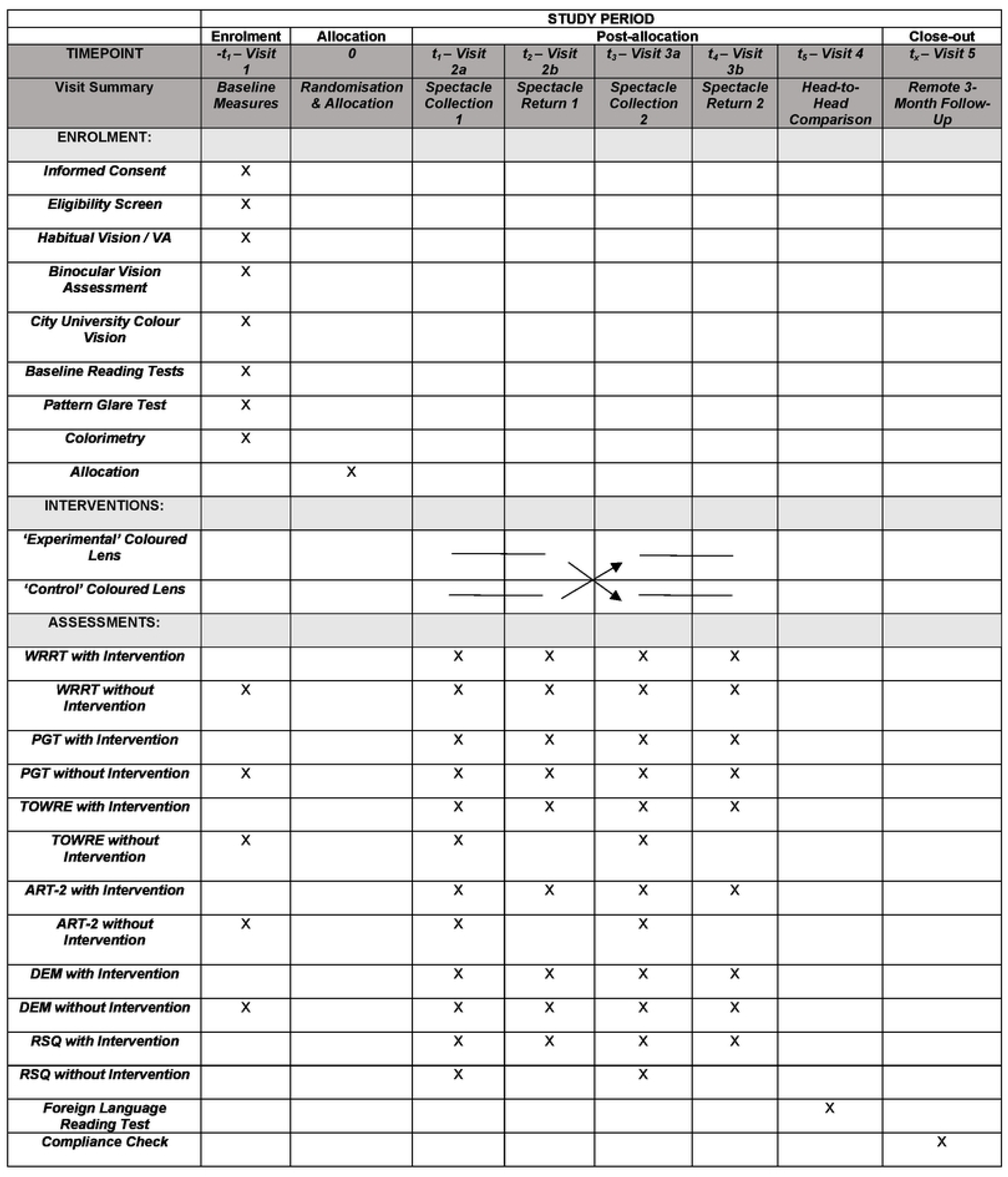
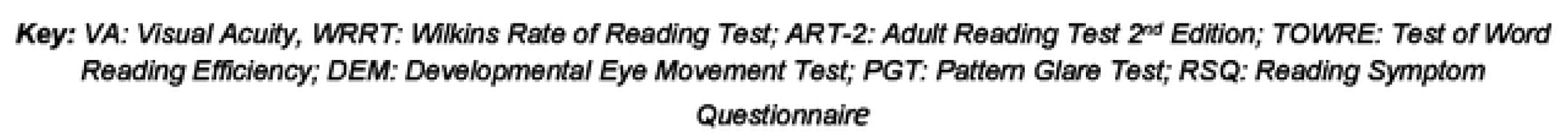
Schedule of Enrolment

**Fig. 2.**
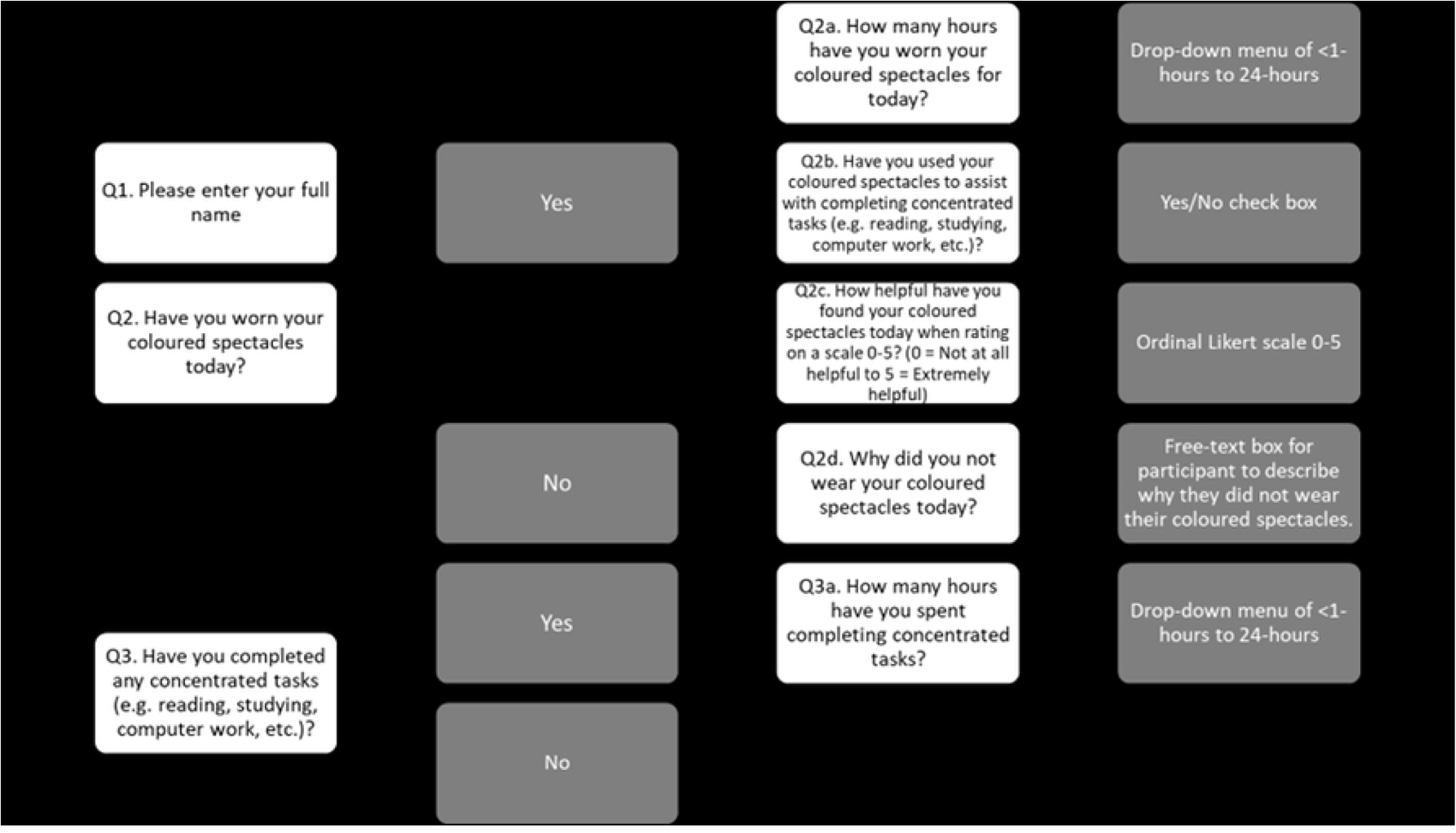
Flowchart of Spectacle Wear Diary Questionnaire

After six weeks of wear, a two week washout period will be initiated. Participants’ spectacles will be collected and stored securely. Washout will ensure potential carryover effects from initial spectacle wear will be eliminated. An overview of trial visits can be seen in the study flowchart in Fig. 3.

**Fig. 3.**
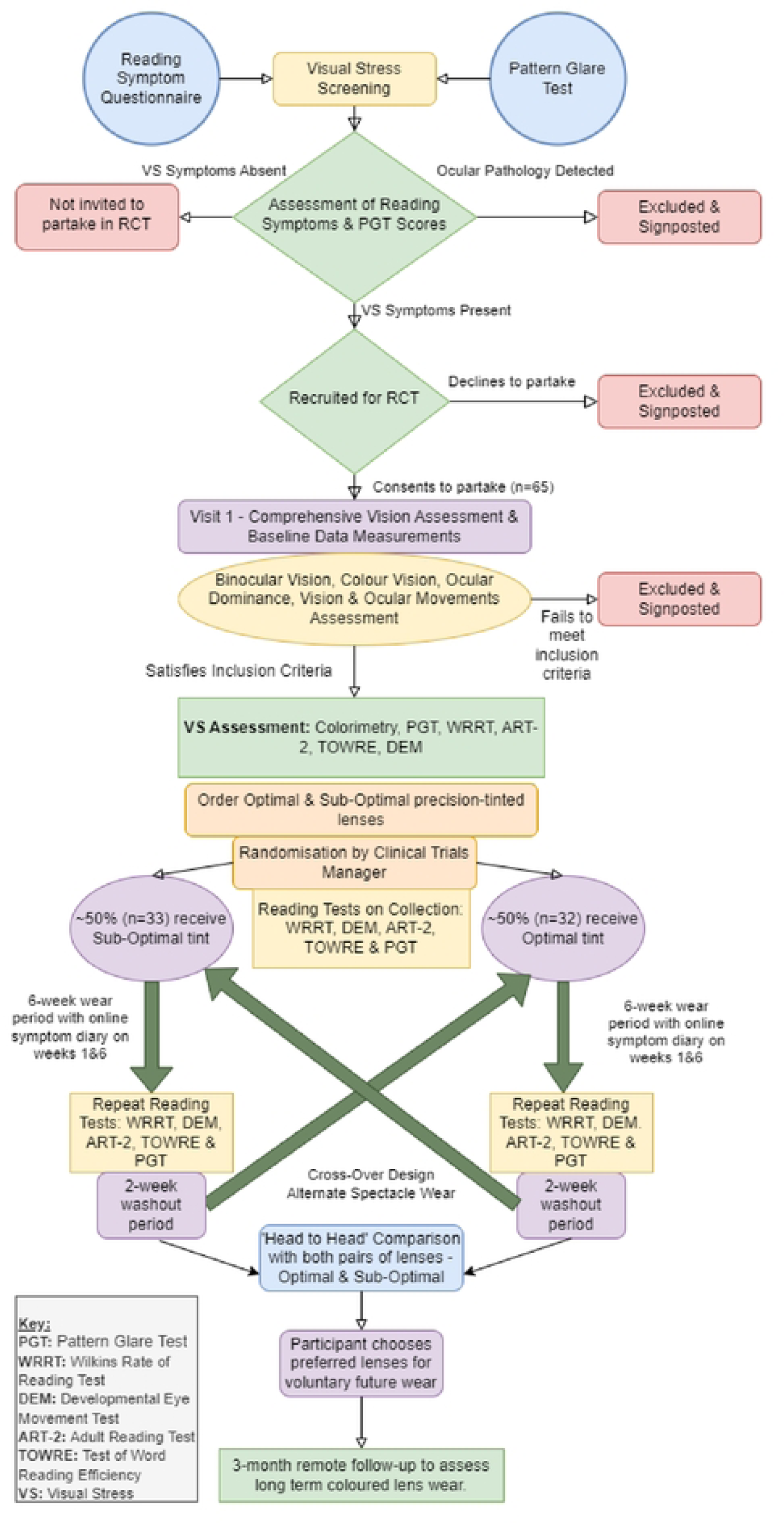
Randomised Control Trial efficacy of coloured lenses for visual stress symptoms Flowchart

Upon completion of wear of both interventions a ‘head-to-head’ comparison of both coloured spectacles will take place. Participants will observe a crowded passage of text in a foreign language with both pair of spectacles under masked conditions. Participants will be asked to consider the reading symptoms they experience and to determine which pair of spectacles they find the greatest perceived benefit, irrespective of measures of reading speed and performance noted earlier in the trial. Participants will be advised to wear the preferred spectacles voluntarily, as they require. Participants will be contacted via telephone after three-months to assess long-term spectacle wear compliance and to assess if a sustained benefit from coloured lenses can be determined. Unmasking of participants shall not take place until the three-month follow-up to avoid influencing compliance levels in long-term spectacle wear.

### Outcome Measures

An overview of the outcome measures employed at each visit during the trial can be seen in Fig. 1 - Schedule of Enrolment, Interventions and Assessment.

#### Primary Outcome Measure: Wilkins Rate of Reading Test Reading Speed

The Wilkins Rate of Reading Test (WRRT) is a long established measure of reading speed with and without the use of coloured filters and has been used in previous reading studies[9][23][24][25][26]. As discussed in full by Wilkins et al. [27], the test comprises 15-randomly-ordered monosyllabic commonly used English language words that repeat throughout 10-lines of a size 9-point text to form a crowded paragraph. Owing to the random order of words presented devoid of context, the test isolates the visual input of processing words whilst reading, so that neither syntactic nor semantic reading processes are involved [52].

Participants will read the passage of crowded text aloud within 60-seconds, with and without coloured lenses as detailed in Fig. 1 – Schedule of Enrolment, throughout the duration of the trial. The number of words-per-minute (wpm) read correctly will be calculated. The examiner will note the number of errors made, namely omissions, reversals, additions, substitutions and mispronunciation [27]. In the event of a ceiling effect whereby all 150 words are read correctly in less than 60 seconds, the time taken to complete the test will be noted, and used to calculate equivalent wpm reading speed [52][53]. The number of additional words read correctly and the percentage reading speed change with coloured lenses will be calculated and compared to baseline measures.

#### Secondary Outcome Measures: Symptom Improvement, Reading Speed with Further Reading Tests, Pattern Glare Scores & Long-term Compliance with Coloured Lenses

##### DEVELOPMENTAL EYE MOVEMENT (DEM) TEST

The Developmental Eye Movement Test (DEM) Version 1 (Bernell Ltd, 1987), as summarised by Facchin [54] and Tanke et al. [55], was originally proposed to measure horizontal saccadic function of readers [56] but is now thought to more broadly assess oculomotor function whilst reading [54]. Horizontal and vertical reading speed will be measured. Number of omission, addition, transposition, and substitution errors will be noted and inform adjusted horizontal and vertical reading speed calculations. A ratio of adjusted horizontal:vertical reading speed will then be determined. The DEM will be completed with and without coloured lenses throughout the trial, owing to the low risk of a learning effect from repeat measures of a test comprising a random series of numbers.

##### TEST OF WORD READING EFFICIENCY (TOWRE)

The Test of Word Reading Efficiency (TOWRE) 1st Edition (Pro-Ed Ltd, Austin TX, 1999) comprises a series of two standardised reading tests which independently assess a participant’s sight-word efficiency and phonemic decoding ability - as discussed further by Torgeson et al. [57]. Reading speed and number of errors made on both subtests will be recorded. A Total Word Efficiency Standard Score will be determined and compared to age-equivalent normative scores and percentiles [57][58]. Reading tests will be completed with and without coloured lenses as detailed in the Schedule of Enrolment.

##### ADULT READING TEST 2ND EDITION (ART-2)

The Adult Reading Test 2nd Edition (ART-2) (Adult Reading Test Ltd, Hayling Island, 2017) will provide participants with a ‘naturalistic’ passage of text to be read aloud [59] with and without the use of coloured lenses, as detailed in the Schedule of Enrolment. The test will measure participants’ comprehension and reading speed under timed conditions. Both ART-2 Test 1 passages – ‘News’ (178 words in two paragraphs; Flesch-Kincaid Grade Level 7.9) and ‘Wildlife’ (179 words in four paragraphs; Flesch-Kincaid Grade Level 7.1), will be alternatively presented to participants to avoid a learning effect. Upon initial collection of ‘experimental’ and ‘control’ lenses, participants will be asked comprehension questions on the completed ART-2 passage, to assess conceptual understanding of the passage. Each set of questions will only be asked once throughout the trial at each spectacle collection appointment, to avoid learning effects.

##### LOW, MID & HIGH SPATIAL FREQUENCY PATTERN GLARE TEST

The Pattern glare test (Institute of Optometry (i.O.O) Sales Ltd, London, UK, 2003) will measure presence and severity of cortical hyperexcitation with and without coloured lenses. Participants will be asked to focus on the horizontal striped patterns of increasing spatial frequencies (0.3, 2.3 and 9.4 cpd) at a fixed 40cm viewing distance as per protocol discussed by Evans & Stevenson [41].

Seven questions relating to perceptual distortion visual symptoms experienced will be posed to participants. Total number of symptoms reported will be summed, where normative values >3 on the mid-spatial frequency grating, may indicate diagnosable pattern glare [41]. The series of pattern glare tests will be repeated throughout the duration of the trial to test the hypothesis that optimally selected coloured filters may reduce severity of pattern glare and thus cortical hyperexcitability.

##### READING SYMPTOM CHANGE

Prior to commencing wear of coloured lenses, participants will be asked to rate reading symptom frequency on the 6-point Likert scale initially used to recruit participants with symptoms suggestive of visual stress to the trial. After 10 minutes and six weeks of coloured lens wear, participants will be asked to repeat the symptom questionnaire. Additionally, after a two week washout period, participants will be asked to rate reading symptom frequency without the use of coloured lenses.

### Statistical Analyses

The Statistical Package for Social Sciences (Version 29.0; IBM Corporation, USA), software will be used for statistical analysis of reading performance and symptom change with use of ‘experimental’ and ‘control’ precision tints.

Average-measures Wilkins Rate of Reading Test reading speed performance with the use of ‘experimental’ and ‘control’ lenses will be assessed by repeat-measures AVOVA testing for both interventions.

Clinically significant improvement noted on the WRRT, will be informed by percentage reading speed improvement with ‘experimental’ lenses, compared to baseline. To date, no validated repeatability values nor defined clinically significant improvement on the test exists in the literature. Therefore, researchers will classify participants’ reading speed improvement by two categories: (i) >5% to 15% improvement; indicating a mild clinical improvement on the WRRT and (ii) >15% improvement; indicating a moderate clinical improvement on the WRRT. These values have been derived from the consensus in the literature, whereby a stricter criterion for reading speed improvement has been applied more recently [3].

Repeat-measures ANOVA testing will also assess reading speed on secondary reading tests employed in the trial with both ‘experimental’ and ‘control’ lenses, for the Adult Reading Test (2^nd^ Edition), The Test of Word Reading Efficiency (TOWRE) and the Developmental Eye Movement (DEM) test. Additionally, repeat-measures ANOVA testing of low, mid and high-spatial frequency pattern glare test scores with both interventions shall be considered. Furthermore, McNemar’s Test of Proportions will investigate changes in reading symptom frequency scores at both the start and end of the six week spectacle wear period, with both spectacle lens interventions when compared to baseline reading symptoms reported.

Additional data incorporated in statistical analysis will include the number of errors made in each reading test with both interventions; number of days spectacle lenses worn; number of hours spectacle lenses worn and number of hours completing concentrated tasks with or without the use of coloured lenses.

### Data Management

Data will be treated and stored in line with the Data Protection Act. Reading symptom questionnaires used will have personal information attached to allow the researchers to identify participants with relevant visual stress symptoms, so that they can be contacted to participate in the trial. Once collected and analysed, each questionnaire shall be anonymised and assigned a unique participant identification number. Participants enrolled in the trial will be identified using their identification number assigned to their reading symptom questionnaire to ensure anonymity during data entry and analysis. All clinical and sensitive information written on hard-copy shall be stored in a secured filing cabinet, of which only the researchers can access. All electronic data will be stored on University-issued encrypted hardware on password-protected and encrypted data analysis programmes. All data (electronic and hard copy) will be held for 10-years in line with university regulations. Anonymised data will be made available upon request upon cessation of the trial.

### Ethical Considerations

The protocol has been approved by the Ulster University, School of Biomedical Sciences Ethical Filter Committee as of 28/10/2022 (Ref: FC-BMS-189). The trial is currently listed on ClinicalTrials.gov (Ref: NCT04318106). Participants will be informed that participation in the trial is voluntary and that they can withdraw their participation at any time. Informed consent will be ascertained through a written consent form provided alongside the reading symptom questionnaire (see supporting information 3 – consent form). Risk of adverse incident from coloured spectacle lens wear is low. In the unlikely event that an adverse incident occurs, it will be reported to the project’s chief investigator S.J.M. and through Ulster University Research Governance protocols.

### Trial Progress To Date

1071 undergraduates at Ulster University have been screened for symptoms of visual stress, 171 undergraduates have met recruitment criteria and have been invited to participate in the trial. 60 participants are currently enrolled in the trial. Researchers plan to screen newly enrolled undergraduate students at the start of the 2024-25 academic year, to ensure adequate trial enrolment.

## Discussion

This study will be the largest randomised control trial investigating coloured lens efficacy in the management of visual stress in adults to date. Previous studies have been criticised for small sample sizes and being underpowered [1], therefore, it has been difficult to draw meaningful conclusions in relation to coloured lens efficacy. Additionally, this randomised control trial will be the first to assess reading symptoms and performance in a homogenous adult population, unlike previous investigations which have included both adults and children [10]. Evidence and guidance in relation to the practice of colorimetry from professional bodies in the United Kingdom is guarded [20][21], advocating for a large scale randomised control trial to take place prior to endorsing the practice. This is echoed in the findings of two recent systematic reviews [1][19]. Therefore, we believe this trial is of high importance to inform best clinical optometric practice.

Recent position statements from the College of Optometrists UK [20] and the Association of Optometrists UK [22] suggest weaknesses in the current evidence surrounding the effectiveness of coloured lenses, and that the treatment of visual stress with coloured lenses remains controversial. Yet, over 300 optometry practices throughout the United Kingdom have an Intuitive Colorimeter™ and assess patients with reading difficulties and symptoms of visual stress, and the wearing of coloured lenses is not uncommon. In addition, optometric practices may offer alternative methods of visual stress assessment, such as coloured overlay assessment, and ‘ChromaGen’ contact lens fitting, amongst others. Whilst this study will not directly influence the management of visual stress with these alternative treatments, further knowledge on coloured filter efficacy will be uncovered from the results of this trial. The results of this study will provide the much needed evidence-base for optometrists, orthoptists and other practitioners assessing visual stress and reading difficulties. Results of this trial may steer the future direction of this area of optometric practice in relation to the practice of colorimetry and prescription of precision tinted lenses.

## Data Availability

Anonymised data will be made available upon request after completion of the trial.

## Acknowledgements

The authors would like to thank the Local Ophthalmic Committee Central Optical Fund for funding the coloured spectacle lenses and spectacle frames. The authors would also like to thank study participants for their commitment to take part in the trial. Additionally, the authors would like to thank technical and auxiliary staff at the Centre for Optometry and Vision Science, Ulster University for their support whilst conducting the trial.

